# Determinants of Maternal Health Service Utilization and Continuum of Care in Nepal: An Analysis from Demographic and Health Survey 2022

**DOI:** 10.1101/2023.08.17.23294226

**Authors:** Achyut Raj Pandey, Bikram Adhikari, Raj Kumar Sangroula, Shophika Regmi, Shreeman Sharma, Bishnu Dulal, Bipul Lamichhane, Saugat Pratap KC, Pratistha Dhakal, Sushil Chandra Baral

## Abstract

**Background:** Continuum of care for maternal health services is essential in minimizing preventable fatalities linked to pregnancy and childbirth. The study focuses on assessing determinants of maternal health service utilization i.e., four or more antenatal care (ANC) visits, institutional delivery, and postnatal care (PNC) visit within the first 2 days of delivery and the continuum of care.

**Methods:** We performed weighted analysis of Nepal Demographic and Health Survey 2022 accounting for complex survey design. Categorical variables are presented using frequency, percentage, and 95% confidence intervals (CI), while numerical variables were represented as mean and a 95% CI. We performed bivariable and multivariable binary logistic regression and the results are odds ratios presented with 95%.

**Results:** Among total participants, 80.5% (95% CI: 77.9, 82.8) had four or more antenatal care (ANC) visits, 79.4% (95% CI: 76.8, 81.9) had institutional delivery and 70.2% (95% CI:67.5, 72.9 postnatal care (PNC) visit within 2 days of delivery. The proportion of participants having both four or more ANC visits and institutional delivery was 67.6% (95% CI: 64.7, 70.4) those completing all three components of care (4 or more ANC visits, delivering in health facility and having PNC visit for mother within 2 days of delivery) was 51.2% (95% CI: 48.3, 54.0).

Compared to participants in poorest wealth quintile, participants in wealthiest quintile had 12 folds higher odds (AOR: 11.96, 95% CI: 14.36, 32.79) of having both four or more ANC visits and institutional delivery. Residents of the Madhesh had lower odds (AOR: 0.47, 95% CI: 0.23, 0.99), Sudurpaschim had higher odds (AOR: 2.37, 95% CI: 1.17, 4.82) of having 4 or more ANC visits and institutional delivery compared to Koshi Province. Residents of Bagmati Province had lower odds (AOR:0.49, 95% CI: 0.28, 0.87) of having all three components of care: 4 or more ANC visits, institutional delivery and PNC visit within 2 days of delivery for mother.

**Conclusion:** There are notable differences in coverage of maternal health services based on education, wealth quintile, province and place of residence. Addressing economic inequalities and provincial differences and harnessing technology to provide and equitable access to vital maternal and newborn health initiatives.

## Introduction

In 2020, an approximate of 800 women tragically lost their lives daily, with one death occurring every two minutes adding up to a total of 287,000 death due to preventable causes associated with pregnancy and childbirth. Of the total maternal deaths occurring worldwide, approximately 95% occurred in low and lower middle-income countries in 2020. There has been notable progress in reducing the global maternal mortality ratio (MMR) over the last two decades, witnessing a decline of around 34% between 2000 and 2020 [1].

Similarly, in 2019, there were approximately 6,700 newborns lost their lives everyday summing up to a total of 2.4 million global deaths among children within the first month of life. In 2019, neonatal mortality accounted for approximately 47% of all child deaths under the age of 5 which is an increase from 40% in 1990 [2], indicating that neonatal period has become the most critical period of interventions. One third of total neonatal deaths occur in the first day of birth and nearly three-quarters within the first week of life [2].

Maternal mortality rate in Nepal stands at 151 per 100,000 live births, with 66% of these deaths occurring in postpartum period, 33% during pregnancy, and 6% during delivery [3]. Between 1990 and 2017, 59,074 more survived pregnancy and childbirth. However, the rate of decline in maternal mortality has been relatively slow in recent years [4]. In 2017, there were approximately 1,364 maternal deaths [4]. For example, between 1990 to 2017, the average annual rate of reduction in maternal materiality ratio was 2.9% while it stood at 0.9 between 2005 to 2017. Hemorrhage is responsible for approximately 15.8% of total maternal deaths in Nepal [4].

In order to achieve the Sustainable Development Goals (SDG) target of 12/1,000 live births, Nepal requires an annual rate of reduction (ARR) in Neonatal Mortality Rate (NMR) of 4.8% which is higher than the current ARR of 4.0% witnessed between year 2000 to 2018, as reported by the Inter-agency Group for Child Mortality Estimation (IGME) [5]. If Nepal achieves SDG target in Nepal there would be 27,116 less newborn deaths 16,434 less stillbirths, 2,208 less maternal deaths, and 5,935 less lifelong disabilities among newborns, compared to maintaining the current coverage of interventions [5].

The causes of most of these maternal and neonatal deaths are preventable [1]. Among the leading causes of maternal deaths are non-obstetric complications (32%), obstetric haemorrhage (26%), hypertensive disorder (12%), pregnancy related infections (7%), self-harm (6%), pregnancy with abortive outcomes (5%) and complication of anesthesia management (1%) [3]. In order to achieve a significant reduction in maternal and neonatal mortality rates and progress towards the elimination of preventable causes of maternal and newborn deaths, it is essential to not only expand the reach of healthcare services but also enhance the overall quality of care provided across the entire spectrum of services [6, 7]. Women who receive midwife-led continuity of care from professional midwives, who are educated and regulated to international standards, experience a 16% lower risk of infant loss and a 24% lower likelihood of preterm birth [2]. These fatalities largely stem from inadequate quality care during pregnancies, childbirth and immediately after birth, as well as during the early days of life [1, 2].

In this context, we aim determinants of four or more antenatal care (ANC), institutional delivery, postnatal care (PNC) within 2 days of delivery, combined coverage of four or more ANC and institutional delivery and also the combined coverage of four or more ANC, institutional delivery and PNC visit within 2 days of delivery

## Methods

This study involves the analysis of data from nationally representative Nepal Demographic and Health Survey conducted between January 5 to June 22, 2022. The sampling frame used for the 2022 NDHS is an updated version of the frame from the 2011 Nepal Population and Housing Census (NPHC), with 36,020 sub-wards which was adjusted for updated urban-rural. The NPHC 2011 there were 58 urban municipalities which has increased to 293 urban municipalities by the time of survey, with these urban areas housing around 65% of the population. The 2022 NDHS adopted an updated urban-rural classification system for the survey.

### Sampling

Two stage stratified sampling technique was used in the study. Seven provinces were divided into urban and rural areas leading to a total of 14 sampling stratum. A strategy of implicit stratification with proportionate allocation was adopted at the lowest administrative levels that included shorting of structuring the sampling frame inside each stratum prior to sample selection, including administrative units from various levels, and using a probability-proportional-to-size strategy during the first sampling step. A total of 476 primary sampling units (PSUs) were chosen using a probability proportional to the size of the PSU of which 248 PSUs were from urban regions, while 228 were from rural areas. A household listing method was carried out in the selected PSUs, yielding a sampling frame. Segmentation was done in sub-wards when the projected number of homes exceeded 300, and only one segment was selected for the survey. The likelihood of selection was related to the size of the segment. Out of the sample, 14,243 households were chosen, and among them, 13,833 were confirmed as being inhabited. From these inhabited households, successful interviews were conducted with 13,786, resulting in a response rate exceeding 99%. All females aged 15 to 49 who were either permanent inhabitants of the designated houses or visitors who had spent the night before in those households were eligible to participate in the interviews. In this manuscript, we have analyzed the data from 1977 women go had live or still births two years preceding the survey.

### Data collection tool

The DHS Program’s model questionnaires were modified to address Nepal’s unique demographics and health problems in consultation with various stakeholders, including government departments, agencies, non-governmental organisations, and funders. After the English versions of the surveys were completed, they were translated into Nepali, Maithili, and Bhojpuri. The translated versions of the questionnaire (household, woman, and man) were then incorporated into tablet computers to facilitate data collection using computer-assisted personal interviews (CAPI). Data for the NDHS 2022 was gathered using 19 teams. Each team comprised of a supervisor, one male interviewer, three female interviewers, and a biomarker specialist.

### Dependent variables

The dependent variables for this study are maternal health service utilization variables, which includes (i) four or more ANC visits, (ii) institutional delivery, and (iii) PNC during the first two days of delivery, (iv) combined coverage of four or more ANC and institutional delivery, and (v) combined coverage of four or more ANC, institutional delivery, and PNC for mother during the first 2 days of delivery.

### Independent variables

Independent variables include ecological belt (mountain, hill, terai), setting (urban, rural), province(Koshi, Madhesh, Bagmati, Gandaki, Lumbini, Karnali, Sudurpaschim), age (in years), ethnicity (Brahmin or Chhetri, Dalit, Janajati, Madhesi, Other), religion (hindu, non-hindu), marital status (unmarried, married or living together, divorced or non-living together), wealth quintile (poorest, poorer, middle, richer, richest), education (no education, basic, secondary, higher), occupation (not working, agriculture, professional or technical or manager or clerical, sales and service, skilled or unskilled labor, others).

### Statistical analysis

We conducted data analysis using R version 4.2.0 and RStudio. To accommodate the complex survey design of NDHS 2022, we employed a weighted analysis approach using the “survey” package. Categorical variables were expressed as frequencies, percentages, and accompanied by a 95% confidence interval (CI), while numerical variables were presented as means along with a 95% CI. To determine the relationship between independent variables and maternal health service utilization variables (including the attainment of at least 4 ANC visits, institutional delivery, and PNC checkup, combined coverage of four or more ANC and institutional delivery, and combined coverage of four or more ANC, institutional delivery and PNC visit within 2 days of delivery), we performed univariate and multivariable logistic regression analyses.

The outcomes of the logistic regression analysis were reported as crude odds ratios and adjusted odds ratios, each accompanied by their respective 95% confidence intervals.

### Ethical approval

The NDHS survey received ethical clearance from Ethical Review Board of Nepal Health Research Council and ICF International. Upon approval of our request for access to data, we downloaded NDHS 2022 dataset from https://www.dhsprogram.com.

## Results

Among total participants, 11% (95% CI: 9.54, 12.7) participants were of age <=20 years, 83.6% (95% CI: 81.7, 85.4) of the age 21-34 years and 5.4% (95% CI: 4.36, 6.62) of the age 35-49 years. The proportion of participants of age <= 20 years was slightly higher in urban 9.6%, 95% CI: 7.68, 11.9) compared to rural (13.8%, 95% CI: 11.6, 16.3) setting. Approximately, 17.7% (95% CI: 15.8, 19.7) of participants at national level, 16.2% (95% CI: 13.8, 19.0) participants in urban and 20.5% (95% CI: 18.0, 23.2) had age <20 years at the time of delivery. Highest proportion of participants were Janajati constituting 30.4% (95% CI: 27.0, 34.1) at national level, 29.2% (95% CI: 24.9, 34.0) at urban areas and 32.7% (95% CI: 27.3, 38.6) at rural areas. More than 80% participants at national level, urban and rural areas were Hindu. Regarding education, highest proportion, 42.9% (95% CI: 39.5, 46.2) of participants had secondary level education followed by 34.0% (95% CI: 31.4, 36.6) who had basic education with similar proportion in urban and rural setting. Slightly less than half of participants, 44.8% (95% CI: 41.3, 48.3) were engaged in agriculture while the husbands of approximately 44.8% (95% CI: 41.3, 48.3) being in the same occupation.

**Table 1:** Characteristics of women aged 15–49 who had a live birth and/or a stillbirth in the 2 years preceding the survey.

|  | **Overall, n = 1,933** |  | **Urban** |  | **Rural** |  |
| --- | --- | --- | --- | --- | --- | --- |
| **Characteristic** | **n** | **% (95% CI)** | **N** | **% (95% CI)** | **n** | **% (95% CI)** |
| Age at delivery |  |  |  |  |  |  |

|  | Overall, n = 1,933 |  | Urban |  | Rural |  |
| --- | --- | --- | --- | --- | --- | --- |
| Characteristic | n | % (95% CI) | N | % (95% CI) | n | % (95% CI) |
| <20years | 342 | 17.7<br>(15.8, 19.7) | 205 | 16.2<br>(13.8, 19.0) | 137 | 20.5<br>(18.0, 23.2) |
| 20-34 years | 1516 | 78.4<br>(76.3, 80.4) | 1011 | 79.8<br>(76.9, 82.5) | 505 | 75.8<br>(73.0, 78.4) |
| >=35 | 75 | 3.9<br>(3.00, 4.98) | 50 | 4<br>(2.86, 5.49) | 24 | 3.7<br>(2.49, 5.40) |
| Ethnicity |  |  |  |  |  |  |
| Brahmin/chhetri | 499 | 25.8<br>(22.8, 29.1) | 337 | 26.6<br>(22.6, 31.1) | 161 | 24.2<br>(20.3, 28.6) |
| Dalit | 359 | 18.5<br>(15.9, 21.6) | 226 | 17.9<br>(14.5, 21.9) | 132 | 19.8<br>(15.9, 24.4) |
| Janajati | 588 | 30.4<br>(27.0, 34.1) | 370 | 29.2<br>(24.9, 34.0) | 218 | 32.7<br>(27.3, 38.6) |
| Madheshi | 354 | 18.3<br>(15.3, 21.7) | 243 | 19.2<br>(15.4, 23.7) | 110 | 16.5<br>(12.3, 22.0) |
| Other | 133 | 6.9<br>(4.46, 10.5) | 89 | 7<br>(4.06, 11.9) | 45 | 6.7<br>(3.22, 13.4) |
| Religion |  |  |  |  |  |  |
| Hindu | 1611 | 83.4<br>(79.9, 86.4) | 1077 | 85.1<br>(80.5, 88.7) | 534 | 80.2<br>(74.4, 84.9) |
| Non-hindu | 321 | 16.6<br>(13.6, 20.1) | 189 | 14.9<br>(11.3, 19.5) | 132 | 19.8<br>(15.1, 25.6) |
| wealth index |  |  |  |  |  |  |
| Poorest | 431 | 22.3<br>(19.7, 25.2) | 177 | 14<br>(11.3, 17.2) | 254 | 38.1<br>(32.8, 43.7) |
| Poorer | 432 | 22.3<br>(19.6, 25.4) | 285 | 22.5<br>(18.9, 26.5) | 147 | 22.1<br>(18.1, 26.6) |
| Middle | 381 | 19.7<br>(17.3, 22.4) | 242 | 19.1<br>(16.0, 22.6) | 139 | 20.8<br>(17.2, 25.0) |
| Richer | 386 | 20<br>(17.5, 22.6) | 283 | 22.4<br>(19.3, 25.8) | 102 | 15.3<br>(11.8, 19.7) |
| Richest | 303 | 15.7<br>(13.1, 18.7) | 279 | 22<br>(18.2, 26.4) | 24 | 3.7<br>(2.45, 5.48) |
| Highest educational level |  |  |  |  |  |  |
| No education | 357 | 18.5<br>(15.9, 21.4) | 221 | 17.5<br>(14.1, 21.4) | 136 | 20.4<br>(16.7, 24.7) |
| Basic | 656 | 34.0<br>(31.4, 36.6) | 403 | 31.8<br>(28.5, 35.3) | 254 | 38.1<br>(34.5, 41.8) |
| Secondary | 828 | 42.9<br>(39.5, 46.2) | 567 | 44.7<br>(40.3, 49.3) | 262 | 39.3<br>(34.8, 43.9) |
| Higher | 91 | 4.7<br>(3.42, 6.48) | 76 | 6<br>(4.14, 8.68) | 15 | 2.3<br>(1.42, 3.58) |
| Participants occupation |  |  |  |  |  |  |
| Agriculture | 866 | 44.8<br>(41.3, 48.3) | 475 | 37.5<br>(33.0, 42.3) | 390 | 58.6<br>(53.8, 63.2) |
| Not working | 801 | 41.4<br>(38.1, 44.8) | 594 | 46.9<br>(42.5, 51.4) | 207 | 31<br>(26.6, 35.9) |
| Professional/technical/managerial /Clerical jobs | 95 | 4.9<br>(3.84, 6.26) | 70 | 5.5<br>(4.05, 7.50) | 25 | 3.7<br>(2.65, 5.25) |
| Characteristic | n | % (95% CI) | N | % (95% CI) | n | % (95% CI) |
| Sales and service | 74 | 3.8<br>(2.93, 5.00) | 55 | 4.3<br>(3.09, 5.99) | 19 | 2.9<br>(1.93, 4.37) |
| Skilled/unskilled labour | 97 | 5<br>(3.93, 6.42) | 72 | 5.7<br>(4.20, 7.70) | 25 | 3.8<br>(2.56, 5.46) |
| Ecological region |  |  |  |  |  |  |
| Mountain | 129 | 6.7<br>(4.56, 9.62) | 50 | 3.9<br>(1.80, 8.41) | 79 | 11.8<br>(8.13, 16.9) |
| Hill | 639 | 33<br>(29.0, 37.3) | 376 | 29.7<br>(24.8, 35.0) | 263 | 39.4<br>(32.7, 46.6) |
| Terai | 1166 | 60.3<br>(56.1, 64.4) | 840 | 66.4<br>(61.0, 71.4) | 325 | 48.8<br>(41.9, 55.7) |
| Province |  |  |  |  |  |  |
| Koshi | 358 | 18.5<br>(16.1, 21.1) | 232 | 18.3<br>(15.4, 21.6) | 126 | 18.8<br>(15.0, 23.4) |
| Madhesh | 500 | 25.8<br>(23.1, 28.8) | 367 | 29<br>(25.4, 32.9) | 132 | 19.9<br>(16.2, 24.1) |
| Bagmati | 295 | 15.3<br>(13.0, 17.9) | 216 | 17.1<br>(14.0, 20.7) | 79 | 11.8<br>(9.17, 15.1) |
| Gandaki | 117 | 6<br>(4.87, 7.46) | 76 | 6<br>(4.47, 8.09) | 40 | 6<br>(4.69, 7.76) |
| Lumbini | 329 | 17<br>(15.2, 19.1) | 185 | 14.6<br>(12.4, 17.1) | 145 | 21.7<br>(18.4, 25.4) |
| Karnali | 149 | 7.7<br>(6.71, 8.86) | 73 | 5.8<br>(4.76, 7.01) | 76 | 11.4<br>(9.35, 13.8) |
| Sudurpashchim | 185 | 9.6<br>(8.36, 11.0) | 117 | 9.2<br>(7.65, 11.0) | 69 | 10.3<br>(8.46, 12.6) |
| Health Insurance Coverage | 215 | 11.1<br>(9.10, 13.6) | 162 | 12.8<br>(9.89, 16.3) | 54 | 8.1<br>(6.16, 10.5) |
| Media exposure |  |  |  |  |  |  |
| No | 1049 | 54.3<br>(51.0, 57.5) | 641 | 50.6<br>(46.2, 55.0) | 408 | 61.2<br>(57.0, 65.2) |
| Yes | 884 | 45.7<br>(42.5, 49.0) | 625 | 49.4<br>(45.0, 53.8) | 259 | 38.8<br>(34.8, 43.0) |
| Internet |  |  |  |  |  |  |
| No | 891 | 46.1<br>(42.8, 49.4) | 498 | 39.3<br>(35.0, 43.8) | 394 | 59.1<br>(54.7, 63.3) |
| Yes | 1041 | 53.9<br>(50.6, 57.2) | 769 | 60.7<br>(56.2, 65.0) | 273 | 40.9<br>(36.7, 45.3) |

Among total participants, 80.5% (95% CI: 77.9, 82.8) participants at national level, 79.5% (95% CI: 75.9, 82.6) participants at urban area and 82.4% (95% CI: 78.9, 85.4) participants in rural area had 4 or more ANC visits. Similarly, 79.4% (95% CI: 76.8, 81.9) participants at national level, 80.9% (95% CI: 77.4, 84.0) in urban and 76.6% (95% CI:72.7, 80.2) rural area had delivery in health facilities. Regarding postnatal care for mothers, 70.2% (95% CI:67.5, 72.9) participants at national level, 71.6% (95% CI: 68.0, 75.0) in urban areas and 67.6% (95% CI:63.3, 71.6) in rural areas had postnatal care visit within 2 days of delivery. The proportion of participants having both four ANC visits and institutional delivery was 67.6 % (95% CI: 64.7, 70.4) at national level, 67.9% (95% CI: 64.1, 71.6) in urban areas, and 67.1% (95% CI: 62.8, 71.2) in rural areas. Further, the proportion of participants completing four ANC visits, delivering in health facility and having PNC visit for mother within 2 days of delivery was 51.2% (95% CI: 48.3, 54.0) at national level, 51.1% (95% CI: 47.4, 54.8) in urban areas and 51.3% (95% CI: 47.0, 55.7) in rural areas.

**Table 2:** Coverage of maternal health service indicators.

| Characteristic | ANC<br>% (95% CI) | ID<br>% (95% CI) | PNC<br>% (95% CI) | ANC+ ID<br>% (95% CI) | ANC+ ID+ PNC<br>% (95% CI) |
| --- | --- | --- | --- | --- | --- |
| Age at birth |  |  |  |  |  |
| <20years | 74.9 (69.6, 79.5) | 80.7 (75.5, 85.0) | 60.7 (54.6, 66.5) | 62.5 (56.8, 67.9) | 45.9 (40.2, 51.8) |
| 20-34 years | 82.2 (79.4, 84.7) | 79.2 (76.3, 81.9) | 63.4 (60.3, 66.4) | 69 (65.7, 72.0) | 52.7 (49.4, 55.9) |
| >=35 | 71.2 (59.2, 80.9) | 78 (66.0, 86.6) | 56.1 (44.4, 67.2) | 64.6 (51.0, 76.1) | 44.8 (32.5, 57.8) |
| Ethnicity |  |  |  |  |  |
| Brahmin/Chhetri | 90.4 (87.4, 92.8) | 87 (83.2, 90.0) | 66.4 (61.7, 70.9) | 79.5 (75.3, 83.1) | 58 (53.3, 62.6) |
| Dalit | 71.4 (64.8, 77.3) | 69.4 (62.6, 75.5) | 56.2 (49.4, 62.7) | 56.1 (49.7, 62.3) | 43.4 (37.1, 50.0) |
| Janajati | 83.9 (80.0, 87.1) | 83.6 (79.1, 87.2) | 66.8 (61.8, 71.6) | 74 (69.0, 78.5) | 57.8 (52.7, 62.8) |
| Madheshi | 72.7 (66.3, 78.2) | 76 (69.5, 81.4) | 57 (50.4, 63.4) | 57.6 (50.9, 63.9) | 40.8 (34.8, 47.1) |
| Other | 73.2 (62.0, 82.0) | 69.3 (55.4, 80.3) | 62 (51.9, 71.2) | 53.1 (39.4, 66.4) | 44.4 (33.2, 56.2) |
| Religion |  |  |  |  |  |
| Hindu | 80.2 (77.5, 82.6) | 79.7 (76.9, 82.3) | 62.7 (59.7, 65.7) | 67.8 (64.7, 70.8) | 51.1 (48.0, 54.1) |
| Non-hindu | 81.9 (75.8, 86.7) | 78 (71.3, 83.4) | 62.2 (55.8, 68.2) | 66.8 (59.7, 73.3) | 51.7 (45.2, 58.2) |
| Wealth index combined |  |  |  |  |  |
| Poorest | 74.5 (69.6, 78.9) | 65.9 (61.0, 70.5) | 50.7 (45.5, 55.9) | 53.5 (48.5, 58.4) | 39.7 (35.1, 44.4) |
| Poorer | 76.7 (71.4, 81.2) | 73.1 (67.5, 78.1) | 59.7 (53.7, 65.5) | 60.5 (55.3, 65.4) | 45.1 (39.5, 50.8) |
| Middle | 77.7 (72.8, 81.9) | 80.1 (75.3, 84.2) | 63 (57.5, 68.1) | 65.4 (59.8, 70.6) | 49 (43.4, 54.6) |
| Richer | 84.5 (79.1, 88.7) | 86.7 (82.0, 90.4) | 67.9 (61.9, 73.3) | 75.9 (70.3, 80.7) | 57.8 (52.1, 63.3) |
| Richest | 92.6 (87.5, 95.8) | 97.6 (94.5, 99.0) | 76.7 (69.9, 82.3) | 90.2 (85.0, 93.8) | 70.4 (63.4, 76.6) |
| Education |  |  |  |  |  |
| No education | 67.2 (60.5, 73.2) | 59.6 (53.5, 65.4) | 50.2 (44.1, 56.3) | 48.4 (42.2, 54.6) | 38.1 (32.5, 44.0) |
| Basic | 75.7 (71.9, 79.1) | 74.2 (69.9, 78.0) | 57.9 (53.3, 62.3) | 58.6 (54.4, 62.6) | 42.6 (38.4, 47.0) |
| Secondary and above | 89 (86.4, 91.2) | 90.9 (88.6, 92.8) | 70.9 (67.3, 74.1) | 81.6 (78.4, 84.5) | 62.3 (58.6, 66.0) |
| Type of place of residence |  |  |  |  |  |
| Urban | 79.5 (75.9, 82.6) | 80.9 (77.4, 84.0) | 62.9 (59.1, 66.5) | 67.9 (64.1, 71.6) | 51.1 (47.4, 54.8) |
| Rural | 82.4 (78.9, 85.4) | 76.6 (72.7, 80.2) | 62.2 (57.9, 66.2) | 67.1 (62.8, 71.2) | 51.3 (47.0, 55.7) |
| Ecological region |  |  |  |  |  |
| Mountain | 90.5 (84.5, 94.3) | 74.9 (61.7, 84.7) | 57.7 (49.1, 65.9) | 70.5 (57.2, 81.1) | 52 (43.1, 60.7) |
| Hill | 86.5 (83.2, 89.2) | 81.8 (78.0, 85.0) | 63.5 (58.5, 68.2) | 74.7 (70.3, 78.6) | 56.2 (51.3, 61.0) |
| Terai | 76 (72.3, 79.5) | 78.7 (74.9, 82.0) | 62.7 (59.0, 66.3) | 63.5 (59.5, 67.3) | 48.4 (44.6, 52.1) |
| Province |  |  |  |  |  |
| Koshi | 78.8 (73.2, 83.4) | 82.2 (76.1, 87.0) | 67.3 (60.3, 73.6) | 66.8 (60.2, 72.8) | 53.4 (46.2, 60.4) |
| Madhesh | 68.4 (61.4, 74.6) | 66.4 (60.6, 71.8) | 52.2 (46.2, 58.1) | 50.1 (44.0, 56.1) | 36.1 (30.5, 42.0) |
| Bagmati | 88.8 (81.8, 93.4) | 88.5 (81.6, 93.0) | 61.4 (52.8, 69.4) | 81 (72.5, 87.3) | 57.5 (48.8, 65.7) |
| Gandaki | 84.6 (77.4, 89.7) | 87.7 (80.4, 92.5) | 71.9 (62.8, 79.5) | 76.3 (68.5, 82.7) | 60.3 (52.5, 67.7) |
| Lumbini | 86.9 (82.1, 90.7) | 84.6 (77.4, 89.7) | 70.3 (64.1, 75.9) | 75.6 (68.8, 81.4) | 60.2 (54.4, 65.6) |
| Karnali | 79.1 (73.7, 83.7) | 71.8 (63.1, 79.2) | 54.8 (46.6, 62.7) | 62.4 (54.2, 70.0) | 45.5 (38.0, 53.1) |
| Sudurpashchim | 90 (85.2, 93.3) | 86.8 (81.4, 90.8) | 70.3 (63.8, 76.2) | 80 (73.8, 85.1) | 60.4 (53.5, 67.0) |
| Covered by health insurance |  |  |  |  |  |
| No | 79 (76.1, 81.6) | 77.7 (74.9, 80.3) | 61.1 (58.1, 64.0) | 65.3 (62.2, 68.3) | 49 (46.1, 52.0) |
| Yes | 92.3 (87.7, 95.3) | 93.4 (88.8, 96.2) | 75.1 (67.2, 81.7) | 86.3 (80.5, 90.6) | 68.3 (60.2, 75.4) |
| Media exposure |  |  |  |  |  |
| No | 77.1 (73.3, 80.4) | 74 (70.3, 77.4) | 59 (55.0, 62.9) | 61.9 (58.0, 65.8) | 47.1 (43.4, 51.0) |
| Yes | 84.5 (81.4, 87.1) | 85.9 (82.9, 88.5) | 67 (63.1, 70.6) | 74.4 (70.8, 77.7) | 56 (52.1, 59.8) |
| Internet |  |  |  |  |  |
| No | 73.5 (69.6, 77.1) | 71.2 (67.4, 74.7) | 57.1 (53.2, 60.9) | 56.7 (52.9, 60.5) | 42.2 (38.6, 45.9) |
| Yes | 86.4 (83.3, 89.0) | 86.5 (83.5, 89.1) | 67.4 (63.5, 71.0) | 77 (73.3, 80.3) | 58.8 (54.7, 62.8) |
| National average | 80.5 (77.9, 82.8) | 79.4% (76.8, 81.9) | 70.2% (67.5, 72.9) | 67.6 % (64.7, 70.4) | 51.2% (48.3, 54.0) |

Compared to participants in poorest wealth quintile, participants in poorer (AOR: 1.77, 95% CI: 1.13, 2.76), middle (AOR: 1.81, 95% CI: 1.16, 2.82), richer (AOR: 2.32, 95% CI: 1.38, 3.90) and richest (AOR: 3.52, 95% CI: 1.66, 7.43) wealth quintile had higher odds of having four or more antenatal care visits. Similarly, participants having secondary or higher education level had two folds higher odds (AOR: 1.58, 95% CI: 1.01, 2.47) of having 4 or more ANC visits compared to participants having no education. Residents of rural area (AOR: 1.37, 95% CI: 1.01, 1.85) had higher odds of having at least 4 ANC visits compared to urban residents while residents of hilly region (AOR: 0.54, 95% CI: 0.31, 0.94) and Terai region (AOR: 0.25, 95% CI: 0.12, 0.49) had lower odds of having 4 or more ANC visits compared to residents in mountain region. Among provinces, residents of Sudurpaschim province (AOR: 2.58, 95% CI: 1.52, 4.38) had 3 folds higher odds of completing 4 ANC visits compared to Koshi Province. Similarly, participants with access to the internet had higher odds (AOR: 1.65, 95% CI: 1.17, 2.31) of having 4 or more ANC visits compared to those who do not have access to the internet.

In comparison to participants aged below 20 years, those within the age range of 20-34 years exhibited diminished likelihood (AOR: 0.62, 95% CI: 0.45, 0.85) of delivering their newborns within a healthcare facility. In contrast, when compared between participants belonging to the most economically disadvantaged wealth quintile, individuals in the poorer (AOR: 1.49, 95% CI: 1.03, 2.16), middle (AOR: 1.86, 95% CI: 1.18, 2.93), richer (AOR: 2.3, 95% CI: 1.28, 4.11), and wealthiest (AOR: 10.23, 95% CI: 3.86, 27.08) quintiles demonstrated elevated odds of delivering their babies within healthcare facilities. Similarly, participants possessing a secondary or higher level of education displayed a twofold increase in the likelihood (AOR: 2.77, 95% CI: 1.87, 4.08) of delivering at healthcare facilities when compared to those without any formal education. Regarding provincial variances, residents of the Madhesh province evidenced reduced odds (AOR: 0.39, 95% CI: 0.22, 0.71), whereas those in Sudurpaschim exhibited a twofold heightened probability (AOR: 2.24, 95% CI: 1.19, 4.20) of delivering at healthcare facilities in comparison to those within the Koshi Province. Correspondingly, participants with internet accessibility demonstrated an increased likelihood (AOR: 1.51, 95% CI: 1.11, 2.05) of undergoing four or more ANC visits compared to their counterparts lacking internet access.

Compared to participants in richest wealth quintile, those in the poorer (AOR: 1.75, 95% CI: 1.22, 2.51), middle (AOR: 1.87, 95% CI: 1.26, 2.78), richer (AOR: 2.16, 95% CI: 1.41, 3.31), and wealthiest (AOR: 3.3, 95% CI: 1.95, 5.60) wealth groups had higher odds of having PNC visit within 2 days of delivery. Regarding province, participants in the Madhesh (AOR: 0.58, 95% CI: 0.35, 0.95) and Bagmati (AOR: 0.56, 95% CI: 0.35, 0.91) had reduced odds of having PNC visit within 2 days.

**Table 3:** Factors associated with four or more ANC visits, ID and PNC within 2 days.

|  | Four or more ANC |  | ID |  | PNC |  |
| --- | --- | --- | --- | --- | --- | --- |
| Characteristic | COR (95% CI) | AOR (95% CI) | COR (95% CI) | AOR (95% CI) | COR (95% CI) | AOR (95% CI) |
| Age at birth |  |  |  |  |  |  |
| <20 years | Ref | Ref | Ref | Ref | Ref | Ref |
| 20-34 years | 1.54 (1.15, 2.07) ** | 1.19 (0.86, 1.64) | 0.85 (0.44, 1.64) | 0.89 (0.44, 1.78) | 1.12 (0.85, 1.47) | 0.91 (0.69, 1.19) |
| >=35 | 0.83 (0.46, 1.51) | 0.79 (0.42, 1.51) | 0.91 (0.66, 1.26) | 0.62 (0.45, 0.85) ** | 0.83 (0.48, 1.42) | 0.75 (0.42, 1.32) |
| Ethnicity |  |  |  |  |  |  |
| Brahmin/Chhetri | Ref | Ref | Ref | Ref | Ref | Ref |
| Dalit | 0.26 (0.17, 0.41) ** | 0.66 (0.40, 1.09) | 0.34 (0.22, 0.52) | 0.88 (0.55, 1.39) | 0.65 (0.46, 0.91) * | 1.09 (0.74, 1.60) |
| Janajati | 0.55 (0.37, 0.83) ** | 0.74 (0.48, 1.16) | 0.76 (0.51, 1.13) | 0.93 (0.58, 1.49) | 1.02 (0.76, 1.37) | 1.27 (0.90, 1.81) |
| Madheshi | 0.28 (0.18, 0.44) ** | 0.7 (0.38, 1.28) | 0.47 (0.30, 0.74) ** | 1.2 (0.63, 2.32) | 0.67 (0.48, 0.94) * | 1.06 (0.64, 1.76) |
| Other | 0.29 (0.16, 0.53) ** | 0.49 (0.20, 1.23) | 0.34 (0.17, 0.66) ** | 0.59 (0.26, 1.35) | 0.82 (0.52, 1.31) | 1.42 (0.71, 2.83) |
| Religion |  |  |  |  |  |  |
| Hindu | Ref | Ref | Ref | Ref | Ref | Ref |
| Non-hindu | 1.12 (0.77, 1.62) | 1.63 (0.96, 2.78) * | 0.9 (0.61, 1.32) | 1.45 (0.90, 2.36) | 0.98 (0.74, 1.30) | 0.95 (0.65, 1.38) |
| Wealth index |  |  |  |  |  |  |
| Poorest | Ref | Ref | Ref | Ref | Ref | Ref |
| Poorer | 1.12 (0.78, 1.61) | 1.77 (1.13, 2.76) * | 1.41 (1.02, 1.94) * | 1.49 (1.03, 2.16) * | 1.44 (1.05, 1.99) * | 1.75 (1.22, 2.51) ** |
| Middle | 1.19 (0.84, 1.69) | 1.81 (1.16, 2.82) ** | 2.09 (1.48, 2.93) ** | 1.86 (1.18, 2.93) ** | 1.65 (1.21, 2.25) ** | 1.87 (1.26, 2.78) ** |
| Richer | 1.87 (1.21, 2.89) ** | 2.32 (1.38, 3.90) ** | 3.38 (2.21, 5.17) ** | 2.3 (1.28, 4.11) ** | 2.06 (1.47, 2.88) ** | 2.16 (1.41, 3.31) ** |
| Richest | 4.3 (2.27, 8.15) | 3.52 (1.66, 7.43) ** | 21.15 (8.80, 50.86) ** | 10.23 (3.86, 27.08) ** | 3.2 (2.13, 4.81) ** | 3.3 (1.95, 5.60) ** |
| Education status |  |  |  |  |  |  |
| No education | Ref | Ref | Ref | Ref | Ref | Ref |
| Basic | 1.52 (1.11, 2.08) ** | 0.96 (0.67, 1.38) | 1.95 (1.45, 2.61) ** | 1.23 (0.91, 1.67) | 1.36 (1.02, 1.82) * | 1.01 (0.72, 1.40) |
| Secondary and above | 3.97 (2.75, 5.73) ** | 1.58 (1.01, 2.47) * | 6.78 (4.74, 9.72) ** | 2.77 (1.87, 4.08) ** | 2.41 (1.80, 3.23) * | 1.44 (0.99, 2.11) |
| Place of residence |  |  |  |  |  |  |
| Urban | Ref | Ref | Ref | Ref | Ref | Ref |
| Rural | 1.21 (0.89, 1.64) | 1.37 (1.01, 1.85) * | 0.77 (0.57, 1.04) | 1.11 (0.82, 1.49) | 0.97 (0.77, 1.23) | 1.21 (0.94, 1.56) |
| Ecological region |  |  |  |  |  |  |
| Mountain | Ref | Ref | Ref | Ref | Ref | Ref |
| Hill | 0.68 (0.36, 1.25) | 0.54 (0.31, 0.94) * | 1.5 (0.77, 2.90) | 1.11 (0.61, 2.01) | 1.27 (0.85, 1.91) | 1.02 (0.71, 1.47) |
| Terai | 0.33 (0.19, 0.60) ** | 0.25 (0.12, 0.49) ** | 1.23 (0.64, 2.37) | 1.73 (0.81, 3.69) | 1.23 (0.84, 1.80) | 0.91 (0.56, 1.47) |
| Province |  |  |  |  |  |  |
| Koshi | Ref | Ref | Ref | Ref | Ref | Ref |
| Madhesh | 0.58 (0.38, 0.90) * | 0.95 (0.55, 1.65) | 0.43 (0.27, 0.67) ** | 0.39 (0.22, 0.71) ** | 0.53 (0.36, 0.78) ** | 0.58 (0.35, 0.95) * |
| Bagmati | 2.14 (1.12, 4.09) * | 0.97 (0.52, 1.84) | 1.66 (0.86, 3.22) | 1.39 (0.71, 2.72) | 0.77 (0.48, 1.23) | 0.56 (0.35, 0.91) * |
| Gandaki | 1.47 (0.84, 2.58) * | 0.96 (0.52, 1.78) | 1.54 (0.79, 3.00) | 1.43 (0.67, 3.08) | 1.24 (0.74, 2.08) | 1.07 (0.62, 1.82) |
| Lumbini | 1.79 (1.11, 2.91) * | 2.11 (1.26, 3.51) ** | 1.19 (0.65, 2.16) | 1.1 (0.61, 1.98) | 1.15 (0.76, 1.74) | 1.11 (0.72, 1.71) |
| Karnali | 1.02 (0.66, 1.57) | 0.79 (0.46, 1.35) | 0.55 (0.32, 0.96) * | 1.04 (0.58, 1.87) | 0.59 (0.38, 0.92) * | 0.8 (0.49, 1.32) |
| Sudurpashchim | 2.42 (1.41, 4.14) ** | 2.58 (1.52, 4.38) ** | 1.42 (0.82, 2.48) | 2.24 (1.19, 4.20) * | 1.15 (0.75, 1.76) | 1.42 (0.88, 2.29) |
| Health insurance coverage |  |  |  |  |  |  |
| No | Ref | Ref | Ref | Ref | Ref | Ref |
| Yes | 3.19 (1.84, 5.55) ** | 1.76 (0.98, 3.16) | 4.08 (2.25, 7.40) ** | 1.66 (0.89, 3.10) | 1.93 (1.28, 2.89) ** | 1.28 (0.83, 1.97) |
| Media exposure |  |  |  |  |  |  |
| No | Ref | Ref | Ref | Ref | Ref | Ref |
| Yes | 1.62 (1.23, 2.14) ** | 1.04 (0.77, 1.40) | 2.14 (1.62, 2.83) ** | 1.15 (0.86, 1.53) | 1.41 (1.12, 1.78) ** | 1.07 (0.83, 1.38) |
| Internet use |  |  |  |  |  |  |
| No | Ref | Ref | Ref | Ref | Ref | Ref |
| Yes | 2.29 (1.70, 3.09) ** | 1.65 (1.17, 2.31) ** | 2.6 (1.98, 3.42) ** | 1.51 (1.11, 2.05) ** | 1.55 (1.23, 1.95) | 1.11 (0.87, 1.40) |

Individuals whose age at the time of childbirth ranged from 20 to 34 years exhibited lower odds (AOR: 0.65, 95% CI: 0.43, 0.98) of undergoing a minimum of four ANC visits and delivering within healthcare facilities. In contrast to the most economically disadvantaged wealth quintile, participants in the poorer (AOR: 1.85, 95% CI: 1.20, 2.86), middle (AOR: 2.4, 95% CI: 1.33, 4.35), richer (AOR: 3.5, 95% CI: 1.82, 6.75), and wealthiest (AOR: 11.96, 95% CI: 14.36, 32.79) wealth quintiles demonstrated elevated odds of experiencing four or more antenatal care visits. Across various provinces, residents of the Madhesh province displayed decreased odds (AOR: 0.47, 95% CI: 0.23, 0.99), while those hailing from Sudurpaschim exhibited twofold higher odds (AOR: 2.37, 95% CI: 1.17, 4.82) of accomplishing the completion of four ANC visits and receiving institutional delivery when compared to their counterparts in the Koshi Province. Similarly, participants with access to the internet exhibited heightened odds (AOR: 1.45, 95% CI: 1.03, 2.04) of undergoing a minimum of four ANC visits and experiencing institutional delivery in comparison to individuals without internet access.

**Table 4:** Factors associated with continuum of care.

| Characteristic | ANC+ID |  | ANC+ID+PNC |  |
| --- | --- | --- | --- | --- |
|  | COR (95% CI) | AOR (95% CI) | COR (95% CI) | AOR (95% CI) |
| Age at birth |  |  |  |  |
| <20years | Ref | Ref | Ref | Ref |
| 20-34 years | 1.045 (0.69, 1.56) | 0.65 (0.43, 0.98) * | 1.17 (0.80, 1.71) | 1.06 (0.72, 1.56) |
| >=35 | 1.92 (0.69, 5.30) | 1.37 (0.49, 3.87) | 0.82 (0.37, 1.80) | 0.67 (0.29, 1.55) |
| Ethnicity |  |  |  |  |
| Brahmin/Chhetri | Ref | Ref | Ref | Ref |
| Dalit | 0.51 (0.32, 0.80) ** | 1.07 (0.65, 1.75) | 1.26 (0.82, 1.95) | 1.42 (0.86, 2.37) |
| Janajati | 1.04 (0.67, 1.61) | 1.28 (0.75, 2.18) | 1.32 (0.91, 1.91) | 1.51 (0.96, 2.39) |
| Madheshi | 0.53 (0.31, 0.89) * | 1.02 (0.48, 2.16) | 0.9 (0.59, 1.38) | 1.05 (0.54, 2.03) |
| Other | 0.37 (0.17, 0.81) * | 0.5 (0.18, 1.37) | 1.87 (0.91, 3.86) | 2.26 (0.82, 6.22) |
| Religion |  |  |  |  |
| Hindu | Ref | Ref | Ref | Ref |
| Non-hindu | 0.81 (0.52, 1.27) | 1.28 (0.74, 2.23) | 1.12 (0.78, 1.61) | 0.81 (0.49, 1.31) |
| Wealth index |  |  |  |  |
| Poorest | Ref | Ref | Ref | Ref |
| Poorer | 1.47 (1.02, 2.11) * | 1.85 (1.20, 2.86) ** | 1.03 (0.66, 1.60) | 1.19 (0.72, 1.97) |
| Middle | 2.09 (1.36, 3.23) ** | 2.4 (1.33, 4.35) ** | 1.04 (0.68, 1.60) | 1.21 (0.69, 2.11) |
| Richer | 3.46 (2.14, 5.57) ** | 3.5 (1.82, 6.75) ** | 1.12 (0.73, 1.70) | 1.32 (0.78, 2.24) |
| Richest | 14.84 (6.17, 35.69) ** | 11.96 (4.36, 32.79) ** | 1.24 (0.79, 1.95) | 1.7 (0.87, 3.36) |
| Education status |  |  |  |  |
| No education | Ref | Ref | Ref | Ref |
| Basic | 1.33 (0.94, 1.88) | 0.75 (0.51, 1.11) | 0.72 (0.44, 1.18) | 0.62 (0.36, 1.09) |
| Secondary and above | 4.27 (2.84, 6.41) ** | 1.58 (0.95, 2.60) | 0.87 (0.57, 1.34) | 0.74 (0.42, 1.30) |
| Place of residence |  |  |  |  |
| Urban | Ref | Ref | Ref | Ref |
| Rural | 0.75 (0.53, 1.05) | 1.14 (0.80, 1.62) | 1.07 (0.80, 1.43) | 1.14 (0.83, 1.56) |
| Ecological region |  |  |  |  |
| Mountain | Ref | Ref | Ref | Ref |
| Hill | 1.78 (0.91, 3.49) | 1.34 (0.73, 2.44) | 1.09 (0.73, 1.62) | 0.96 (0.64, 1.45) |
| Terai | 1.43 (0.73, 2.78) | 1.53 (0.69, 3.43) | 1.14 (0.79, 1.66) | 0.78 (0.45, 1.34) |
| Province |  |  |  |  |
| Koshi | Ref | Ref | Ref | Ref |
| Madhesh | 0.49 (0.29, 0.82) ** | 0.47 (0.23, 0.99) * | 0.65 (0.39, 1.07) | 0.64 (0.33, 1.23) |
| Bagmati | 1.84 (0.86, 3.95) | 1.18 (0.55, 2.53) | 0.61 (0.37, 1.03) | 0.49 (0.28, 0.87) * |
| Gandaki | 1.65 (0.80, 3.43) | 1.25 (0.53, 2.93) | 0.95 (0.50, 1.79) | 0.83 (0.42, 1.65) |
| Lumbini | 1.19 (0.60, 2.38) | 1.13 (0.57, 2.23) | 0.98 (0.59, 1.62) | 0.96 (0.55, 1.67) |
| Karnali | 0.67 (0.36, 1.24) | 1.28 (0.66, 2.50) | 0.67 (0.38, 1.19) | 0.72 (0.38, 1.38) |
| Sudurpashchim | 1.44 (0.79, 2.63) | 2.37 (1.17, 4.82) * | 0.77 (0.45, 1.33) | 0.86 (0.47, 1.55) |
| Health insurance coverage |  |  |  |  |
| No | Ref | Ref | Ref | Ref |
| Yes | 3.01 (1.61, 5.64) ** | 1.35 (0.71, 2.60) | 1.26 (0.79, 2.00) | 1.14 (0.70, 1.84) |
| Media exposure |  |  |  |  |
| No | Ref | Ref | Ref | Ref |
| Yes | 1.81 (1.33, 2.46) ** | 1.03 (0.74, 1.43) | 0.95 (0.73, 1.25) | 0.93 (0.69, 1.26) |
| Internet use |  |  |  |  |
| No | Ref | Ref | Ref | Ref |
| Yes | 2.42 (1.75, 3.34) ** | 1.45 (1.03, 2.04) * | 1.11 (0.83, 1.49) | 1.1 (0.79, 1.52) |

Residents of Bagmati Province had lower odds (AOR:0.49, 95% CI: 0.28, 0.87) of having all three components of care: 4 or more ANC visits, institutional delivery and PNC visit within 2 days of delivery for mother.

## Discussion

For women’s reproductive health, it is essential to provide continuous maternal health care. In the context of MCH, care provided from the time of conception through childbirth, the postnatal period, infancy, and the childhood period is referred to as Continuum of Care [8, 9]. If a woman uses all three services as part of a continuum of maternity care, the government of Nepal has a program that offers all three services free of charge to customers. The aim of this study was to assess the status of continuum of care and explore the factors associated with continuum of care for MNCH care.

### ANC

Among the total participants, at least 80% of women had four ANC visits in the 2 years preceding the survey which was greater than the four or more ANC coverages in years 70.8% in 2016 [10] and 53.1% in 2011 [10] and 77.9% in 2019 [11]. The four or more ANC coverage in Nepal is higher than the recent DHS conducted in South-East Asian countries like India where it stood at 57.9% in 2019-21 and Bangladesh where it stood at 45.8 % in 2017/18, 52.8% in Pakistan in 2017-18 [10]. In this study, the women residing in rural settings had higher odds of receiving at least four ANC compared to the women residing in urban settings. The result of this study contradicts with the findings from a further analysis of NDHS 2016 [12], 2019–2021 National Family Health Survey, India [13] and a meta-analysis of Demographic and Health Survey Data conducted which 28 low- or middle-income countries [14]. In 2016, the coverage of 4 or more ANC visits was 75.5% in urban and 61.7% in rural setting with relatively higher coverage in urban areas [15]. However, the situation reversed in 2022, with coverage of 4 or more ANC visits standing at 79.5% in urban and 69.2% in rural areas [16]. The reason for this reversal in coverage status in urban and rural settings is not clearly understood. However, after 2015, Nepal had massive changes in health system with three tiers of governments being functional and taking responsibility of basic health services including maternal health services. The transfer of authority to local level governments may have led to relatively better planning and effective implementation of interventions which increased the coverage in rural areas at a faster pace than urban areas thus surpassing the coverage figures. In Nepal, over 51,000 active FCHVs play a primary role in advocating for safe motherhood and encouraging the utilization of newborn health services, demonstrating notable outreach, particularly in rural areas.

After transition to federal structures, local governments have sought to have more active role of FCHVs in identification of households with pregnant women and thus counselling them for service utilization. This could have further accelerated the coverage of 4 or more ANC in rural settings.

As compared to the women residing in hill regions, women residing in mountain regions have higher odds and people residing in terai have lower odds of having 4 or more ANC. In one of the previous article, that involved further analysis of NDHS 2016, 4 or more ANC visit was found to be 68.44% in mountain, 76.63% in hill and 66.14% in terai although no statistically significant association was revealed in multivariable analysis [12]. In 2022, the coverage of 4 or more ANC visits stands at 90.5% in Mountain, 86.5% in Hill and 76% in Terai belt [15]. Comparable to changes in indicators in urban/rural setting, there seems to be relatively faster progress in mountain regions compared to hilly and terai regions. Similar to urban/rural changes, transition of health system to federal structure after 2016 with transfer of authority to local and provincial governments may have created additional space for more contextualized planning.

Women residing in Lumbini and Sudurpaschim province had higher odds of receiving four or more ANC as compared to the women residing in Koshi province. Variations in general development may be partially responsible for regional differences in ANC visits, and these differences also lead to gaps in healthcare access.

Our findings show significant differences in antenatal visit across maternal educational attainment and wealth index. Women from the poorest households and those without formal education were most at risk for receiving insufficient ANC. This is in line with the further analysis of the recent National Family Health Survey, India [13, 17]. The further analysis of 2016 DHS in Nepal and 2017 DHS, Indonesia also has similar findings [12, 18]. The women using internet had higher odds of receiving four or more ANC visits in this study. Improved access to the internet is directly proportional to healthcare service accessibility. It affects socioeconomic determinants of health like education, employment, and access to healthcare that are more widely known to play a role in health care outcomes [19].

### Institutional Delivery

Institutional delivery in Nepal according to NDHS in the 2 years preceding the survey was 79% which was higher than the previous two survey conducted: 57% in 2016 and 35% in 2011 [20]. The NMICS survey conducted in 2019 had slightly lower coverage of institutional delivery than this study which was 77.5% [11]. Nepal has a greater institutional delivery coverage rate than recent DHS surveys in South-East Asian nations like Bangladesh, where the rate was 65% [21], and in Pakistan, where the rate was 66% [22]. However, the institutional delivery in this study was lower than the recent national-level demographic survey conducted in India which was 89% [23].

In this study, the odds of giving birth at a health facility was lower in Terai region as compared to hill region. The odds of delivering in a health facility was lower in Madhesh province and higher in Sudurpaschim province as compared to Koshi Province which is supported by further analysis of multi-indicator cluster survey, 2019 [24]. There may be differences in the availability and use of maternal health services among the provinces that contribute to the uneven use of birthing facilities among the different provinces.

Among the age categories of women delivering in health facilities, women belonging to 20-34 years age group had lower odds of institutional delivery as compared to the women of age less than 20 years. Further analyses of demographic and health survey conducted in different countries support the findings of this study [25–28].

Similar to other studies conducted in Nepal [24, 29], this study also identified that women of higher wealth quintile were more likely than women with lower wealth quintile to give birth in a health facility. Other studies conducted in Ethiopia [27], India [25], and Zambia [28] found that the household wealth index was positively associated with institutional delivery.

In this study, women with higher education had higher odds of delivering at heath facility than the women with no education or lower education and this finding is in line with the other studies conducted in Nepal [24, 29] and other parts of the world [25–27, 30]. Women who have more education may be able to make decisions about giving birth in a health facility. Internet use was positively associated with institutional delivery in this study and is supported by the further analysis of NMICS 2019 [24]. Women using the internet may be exposed to different information regarding maternal and child health services through different social medias which have contributed to increased institutional delivery.

### PNC

Among the total participants in this study, within the first two days following delivery, 70% of women and newborns received a postnatal check. In the previous DHS survey conducted in 2016, 57% of women reported having received a PNC checkup in the first 2 days after birth and in DHS 2011 it was 45% [20]. The proportion of women who received PNC check up in the first two days of delivery was 69.4% in the MICS survey 2019 [11]. The recent national demographic surveys conducted in Pakistan and Bangladesh has lower coverage of postnatal checkup with only 60% in Pakistan [22] and 55% in Bangladesh [21]. However, according to the national level demographic study conducted in India, the proportion of women who received PNC check up in the first two days of delivery was 82% [23] which is higher than this study.

In this study, Madhesh and Bagmati province had lower odds of institutional delivery as compared to Koshi province. The exact reason for these differences is not known and may require further qualitative studies. However, management approaches and strategies adopted in expanding coverage within the provinces could be responsible for differences.

This study identified that women with higher wealth quintile were more likely to conduct PNC checkup within 2 days of delivery than women with lower wealth quintile. This finding is in line with the studies conducted in Nepal [31] and Ethiopia [32]. Women with high financial standing may have the freedom to decide on the use of household incomes. They may also be able to pay for the costs of their own and their children’s health care.

Women with secondary or higher education are more likely to receive early PNC as compared to women with no education. This result is in accord with recent studies showing that higher education was significantly associated with postnatal care attendance among women [9, 33, 34]. Regarding occupation of the women, women who were skilled or unskilled labour were more likely to receive postnatal checkup than the women with agriculture as occupation.

Women who had health insurance were more likely to receive postnatal checkup than the women who did not have health insurance and this is in line with the study conducted in Sub-Saharan Africa [9]. Health insurance acts as an intervention that enables individuals to complete the Continuum of Care with little to no out-of-pocket expense.

### Wealth quintile and continuum of care

In our study, compared to poorest quintile, participants in poorer, middle, richer and richest wealth quintile were found to have higher odds of completing both the four or more ANC visits and delivering in health facilities which also aligns with some of the previous studies [35, 36]. Previous studies suggest that the most effective method for making rapid advancements in delivering Maternal and Child Health services involves extending support to socio-economically disadvantaged communities. Despite the availability of free maternal health services and incentives for service utilization, our study indicates that economic disparities play a role in the uneven utilization of healthcare services. The expansion of community-based services and awareness campaigns, bolstering primary healthcare facilities, and implementation of other measures targeting the economically marginalized are of utmost importance. However, combining all three variables in a composite index (4 or more ANC visits, ID and PNC visits within 2 days) was not found to be associated with wealth quintile. The reason for this difference is not clearly understood and further studies could be useful in this area.

### Provincial variation

Provincial variations were noted in continuum of care for maternal health services utilization with residents of Sudurpaschim having two folds higher odds of having 4+ ANC visits and institutional delivery. These variations based on administrative divisions and geographic area were also reported in multiple previous studies in different settings like India [36], Pakistan [35] and Ethiopia [36]. Differences based on provinces and geography could be because of differences in programme implementation strategies adopted, management efficiency and socio-cultural variation among provinces.

### Education and continuum of care

Our study revealed that women having secondary level or higher education are more likely to complete four or more ANC visit and deliver in health facilities. Multiple previous studies have revealed the association of education level with continuum of care [35, 37–39]. This could be because education may improve women’s understanding, ability to access information.This could be attributed to education as education can help enhance women’s comprehension, facilitate their information access and enable a more thorough understanding of advocacy messages communicated through media, the internet, and healthcare providers. Additionally, women might have better access to sources of information and possess more knowledge about the services and feel more confident in service utilization which cumulatively result in better service utilization.

### Age and continuum of care

Participants of age 20-34 years seem to have lower odds of completing 4 or more ANC visits and institutional delivery. Participants of this age group have lower odds of utilizing institutional delivery services, while the findings were not significant for 4 or more ANC visits and Institutional delivery services.

### Internet use

Participants using the internet were found to have slightly higher odds of maintaining the continuum of care till institutional delivery completing 4 or more ANC visits and delivering in health facilities. Use of the internet increases access to health educational messages which could be one factor for completing both 4 or more ANC visits and institutional delivery.

### Policy Implication

While Nepal government has made significant progress in improving maternal health services line ANC visits, institutional delivery and PNC visits, additional efforts are needed to ensure that the chain is maintained across continuum of care. Counselling during 1^st^ ANC visits for the follow up visits, and for institutional delivery could be useful strategy which is already reflected in policies of MoHP. Ensuring quality of care and gaining trust would require regular supervision and quality assurance activities at facility level.

### Strengths and Limitations

Our finding are based on further analysis from nationally representative data collected in globally standardized tool, taking into account the recent federal structure during samplings process. As the survey has used standardized definition for variables, and has used globally accepted survey methodology, findings are comparable with findings from other countries. However, as the survey was undertaken while COVID-19 pandemic was ongoing, there could be some over or underestimation in case of some variables.

## Conclusions

Several factors influenced maternal health service utilization, including wealth quintile, education level, place of residence, and access to the internet. Addressing these disparities is imperative to ensure that all strata of the population can benefit equally from improved maternal and newborn health services. In addition, by aligning efforts across the continuum of care, Nepal can accelerate progress toward its maternal and newborn health goals, reducing preventable deaths and ensuring the well-being of mothers and infants.

## Data Availability

Data are available for download on request from: https://www.dhsprogram.com.

